## Supplemental Material for "Deep Optical Blood Analysis: COVID-19 Detection as a Case Study in Next Generation Blood Screening"

### 1. Model Training and Architecture

#### *1.1. Training Infrastructure*

To implement the neural network models used throughout this work we used PyTorch [9]. During the training process we applied data augmentation to individual blood cell images across both branches and their respective training regimes. The images were augmented using *Torchvision* [8] with standard horizontal/vertical flipping, random rotations, and color jittering. We tracked performance on the held-out validation set during model training using *Weights & Biases*[2], with the final model used for evaluation being the iteration of the model which performed best on the validation set.

To test our system across all available data, we used repeated stratified k-fold cross validation from *Scikit-Learn* [10] on a patient level based their COVID-19 test result. There were a total of 236 patients, 125 which were labelled positive for COVID-19. We first split our data into a 83%-17% training/testing split, then split the training data again with a 80%-20% split for our final training and validation sets. The proportion of COVID-19 positive patients was approximately balanced within each set, and there was no cross-over of patient data from set-to-set. We rotated through our dataset, for a total of six folds (a single fold consisting of a unique training/validation/test set). Unless otherwise indicated, all performance metrics are reported as the average test-set performance across all six folds, where multiple independent models were trained from scratch exclusively on the individual folds. This strategy enabled us to test our system on all available data while isolating the test data during the training process.

#### *1.2. Single-Image Model*

The single-image branch of our hybrid deep learning system was configured to have two outputs for each image, representing the probabilities of the image being either COVID-19 positive or negative. We used the DenseNet-121 [4] neural network architecture provided by

PyTorch [9], which has a $240 \times 240 \times 3$ input size. To optimize the model we chose the stochastic gradient descent optimizer included in PyTorch [9] with standard cross-entropy loss, paired with a cyclic learning rate schedule (following Smith [11]), linearly oscillating between high and low learning rates ($1$ and $0.0001$ respectively) every $4000$ gradient steps. The *BatchNorm* [5] layers within the DenseNet-121 architecture allowed for high learning rates (e.g. $1$) without harming optimization. Empirically we found that when trained from random initialization, the *DenseNet-121* model performed better than its pre-trained alternatives.

Our single-image dataset was constructed by propagating labels from the patient to all individual images (for example if a patient was COVID-19 positive then all of their data would be labelled as COVID-19 positive). During training we used a weighted random sampling strategy to sample an equal amount of COVID-19 positive and negative images from our dataset.

The single-image model was trained for a total of $800$ epochs. To generate validation metrics representative of our task we evaluated our model by taking the average of all outputs across a patient's data (i.e. running every image included within a patient's PBS and taking the average output as the patient prediction). The final model selected was the model which had peak validation performance, which is evaluated at the end of every epoch.

#### *1.3. Multi-Image Model*

The multi-image branch of our system consists of three steps: feature extraction, attention based aggregation, and classification. For the feature extraction portion of our pipeline we chose the ResNet50 [3] neural network architecture, configured to produce feature vectors, $f_i \in \mathcal{R}^{32}$, for each image $i$. The attention mechanism used a small multilayer perceptron (MLP) with one hidden layer of size $64$ and a *tanh* activation to produce a single *importance* value per image ($a_i$). Each importance value is placed into a softmax function to produce a set of feature weights $W$, such that $w_i = \frac{e^{a_i}}{\sum_{j=1}^{N} e^{a_j}}$ and $\sum_{j=1}^{N} w_j = 1$. These weights are used in conjunction with the extracted features ($f_i$) to create a final per-image set feature vector $F = \sum_{j=1}^{N} f_j w_j$. Finally this feature vector is transformed into a classification with a single linear layer.

Several components of our multi-image branch were critical to successful performance. The most important component was the use of a unique random image subset generation process for model training, which was not used during inference. Specifically, during model training, we randomly selected a subset of 16 images per patient (from all available per-patient images) to form our model input. This random set of 16 training images changed at each iteration step of model optimization. During model inference, however, we used all available images from each patient (see Figure 2 for the distribution of image count across patients). This random sampling strategy first helped address high memory usage encountered during network training, Second, we hypothesize that it also had the beneficial effect of acting as a regularization strategy (akin to image augmentation), preventing the network from utilizing relatively few images within a patient's PBS image set to drive predictions.

We trained our multi-image model for $2000$ epochs. Similar to the single-image model validation performance was evaluated by examining the prediction of the patient's infections status using *all* of their data, and the model weights which achieved peak performance were saved for final evaluation. Experimentally, we found that the multi-image branch was much more sensitive to learning rate, with large learning rates causing divergence. To accommodate this sensitivity, we modified our training strategy from the single image branch and used an adaptive optimizer (AdamW [6]) with a small, fixed, learning rate ($3e - 5$).

| Parameter | Single-Image | Multi-Image |
|---|---|---|
| CNN Architecture | DenseNet-121 | ResNet50 |
| Minimum Learning Rate | $1 \times 10^{-5}$ | $3 \times 10^{-5}$ |
| Maximum Learning Rate | 1 | $3 \times 10^{-5}$ |
| Optimizer | SGD | AdamW [6] |
| Batch Size | 32 | 8 |
| Images per Sample | 1 | 16 |

Table 1: Model Training and Configuration Parameters

### 2. Data Preprocessing and Augmentation

We applied the same kind of data preprocessing and augmentations to all images used within this study. To prepare the data for processing by our neural networks we cropped and normalized the images. The cropping was done by taking the inner $240 \times 240$px from the original $360 \times 360$px image. We normalized the images by calculating a dataset wide mean and standard deviation for each color channel (red, blue, and green), then subtracting the mean and dividing by the standard deviation.

To augment our dataset we applied, in order, random horizontal and vertical flipping, random rotation, and color jittering. These augmentations were implemented using TorchVision [8] with parameters reported in Table 2. The aforementioned cropping step was done *after* the augmentations had been applied. During our perturbation experiments we added a final masking step to our augmentation pipeline, replacing content within the mask area with zeros.

| Parameter | Value |
|---|---|
| Random Horizontal Flipping | $50\%$ probability |
| Random Vertical Flipping | $50\%$ probability |
| Color Jitter | $\pm[0, 5]\%$ Brightness<br>$\pm[0, 5]\%$ Contrast<br>$\pm[0, 5]\%$ Saturation<br>$\pm[0, 5]\%$ Hue |
| Random Rotation | Uniformly sampled from $[-45, 45]$ |
| Center Crop | Center $240 \times 240$px |

Table 2: Data augmentation and pre-processing parameters (see Torchvision [8] for implementation details). Transformations are applied in the order they appear within the table. The same transformations were applied in both the single and multi-image models.

### 3. White blood cell classifier

Within the main body of this work we drew relationships between the attention values produced by one of our multiple instance learning (MIL) methods and categories of white blood cells (WBCs). To develop this relationship we first needed to be able to classify each of the individual cells into predefined categories. To enable WBC classification we used a pre-existing open-source dataset produced by Acevedo et al. [1], which used the same model of imaging system (*Cellavision DM9600*) used within our work. Within their dataset they classified WBCs into eight distinct classes: Neutrophils, Eosiophils, Basophils, Lymphocytes, Monocytes, Immature Granulocytes (IG), Erythroblasts, and Platelets. However, we found that within our dataset there were a large number of what are canonically known as *Smudged Cells*, these are WBCs which are broken during the slide making process [7], these cells were not represented within the dataset provided by Acevedo et al. [1]. To compensate for this exclusion we manually labelled 2000 images within our main dataset and included them with the 17,000 images of the other classes.

Using this expanded dataset we trained a convolutional neural network (CNN) to predict the classification of each cell. We followed a similar procedure to the single-image classifier used for detecting COVID-19, using the same set of image augmentations and the ResNet50 [3] network architecture, trained from random initialization. We used the AdamW optimizer [6] with a fixed learning rate of $3\times10^{-4}$, and trained the model for $100$ epochs. We used stratified K-Fold cross validation to ensure equal proportions of each cell type across our training, validation, and test sets of $70\%$, $15\%$, and $15\%$ respectively. With this training strategy our model was able to achieve an average classification accuracy across our test set of $92\%$

### 4. Scan Data Statistics

To understand the underlying trends within our data we plotted the distribution of WBC types (determined by our automated WBC classifier). Within Figure 1 we can see that the most common cell is the neutrophil, constituting $\sim 35\%$ of the images, while other cell types occur at a medium to low frequency. We also examined the number of images per-patient, as shown in Figure 2. Here we can see that there is no meaningful difference between image counts between condition (COVID-19 positive vs. negative), however there is a great deal of variation across the entire dataset.

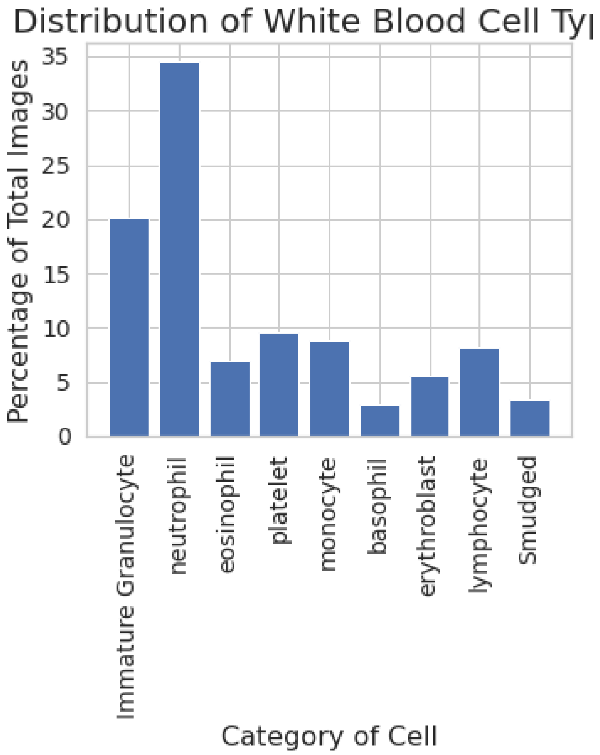


Figure 1: Distribution of images by white blood cell classification (using automated WBC classifier).

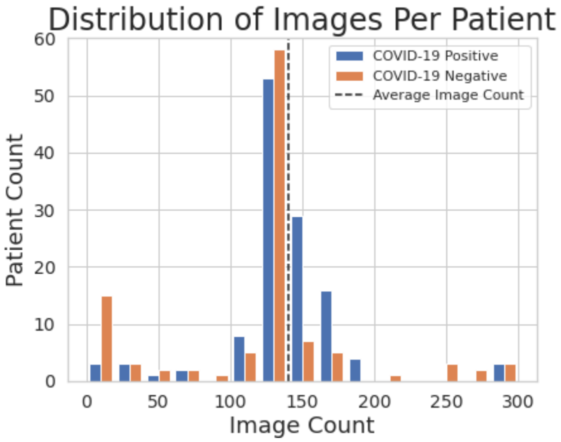


Figure 2: Number of images per patient across the entire standard dataset. There was a large degree of variance between patients in image count, with an average of 134 images per patient.

[2] Lukas Biewald. Experiment tracking with weights and biases, 2020. Software available from wandb.com.

[3] Kaiming He, Xiangyu Zhang, Shaoqing Ren, and Jian Sun. Deep residual learning for image recognition. In *Proceedings of the IEEE conference on computer vision and pattern recognition*, pages 770–778, 2016.

[4] Gao Huang, Zhuang Liu, Laurens Van Der Maaten, and Kilian Q Weinberger. Densely connected convolutional networks. In *Proceedings of the IEEE conference on computer vision and pattern recognition*, pages 4700–4708, 2017.

[5] Sergey Ioffe and Christian Szegedy. Batch normalization: Accelerating deep network training by reducing internal covariate shift. *arXiv preprint arXiv:1502.03167*, 2015.

[6] Ilya Loshchilov and Frank Hutter. Decoupled weight decay regularization. *arXiv preprint arXiv:1711.05101*, 2017.
